# Current attitudes towards HIV viral load self-testing: thematic analysis of a consultation with potential end-users

**DOI:** 10.1101/2021.08.27.21260566

**Authors:** Harriet D Gliddon, Rachel A McKendry, Jo Gibbs

## Abstract

**Background:** HIV viral load (VL) is key to monitoring response to antiretroviral therapy. A number of point-of-care HIV VL tests are in development. These tests could be repurposed for HIV VL self-testing.

**Methods:** We held a patient and public involvement (PPI) consultation for people living with HIV to explore the prospect of HIV viral load self-testing. We conducted a thematic analysis on the data from this consultation.

**Results:** Key themes were access, convenience, usability, technical aspects, technology potential, connectivity and confidentiality. Attendees expressed significant appetite for decentralised HIV care, including HIV VL self-testing.

**Conclusions:** This PPI consultation can help researchers developing HIV viral load self-testing technology to better understand the needs of potential end users. Our findings will help guide the development and implementation of HIV VL self-testing.

## Introduction

The World Health Organization (WHO) recommends HIV VL testing as the most effective means of monitoring antiretroviral treatment (ART) success^1^. In the UK, guidelines recommend HIV VL testing every six to 12 months for people established on ART and with a suppressed VL^2^. These appointments often involve other routine monitoring including organ function tests and full blood count. People living with HIV have emphasised the importance of building a strong relationship with their clinician, and the reassurance it brings^3^. However, busy HIV outpatient clinics and limited consultation times have negatively affected continuity of care^3^. Appointments often require individuals to take time off work for clinic attendance^4^ and travel relatively long distances^5^. Shorter waiting times for appointments and appointments outside of working hours are important to HIV patients^6^ and measures to avoid inconvenience should be taken to improve retention in care and ART adherence^7^. The COVID-19 pandemic has increased the need for measures that facilitate remote consultations and increase time periods between monitoring visits. New service models have evolved and remote consultations for primary care and outpatient appointments have become accepted as a new norm^8^. There is now a clear precedent for technology that facilitates these remote consultations for a number of health conditions, including HIV.

A number of point-of-care (POC) HIV VL tests have been designed for use in resource-limited settings, where laboratory infrastructure is not available^9^. New devices are now able to provide near-patient VL testing and offer improved HIV management through shorter turnaround times for results. A recent study conducted in Durban, South Africa, showed that POC HIV VL testing improved viral suppression and retention in HIV care^10^. There is clear potential for these to be used in outreach settings in other health systems, including the UK. It is also possible that these POC HIV VL tests could be repurposed as HIV VL self-tests, for people living with HIV to administer themselves. Although self-testing for HIV was little thought of when HIV POC tests were first developed, HIV self-tests now form an important component in the prevention toolbox for HIV. HIV self-tests were licensed in the UK in 2014, have been shown to be acceptable in high-risk populations, and can improve testing coverage among those who have not tested previously^11 12^. In 2019, WHO released guidelines on self-care interventions with a focus on sexual and reproductive health and rights^13^. These guidelines strongly recommended the provision of both HIV self-tests and interventions as a means of imparting self-efficacy, knowledge and empowerment for people living with HIV. Given the success of repurposing POC HIV tests as HIV self-tests, it is pertinent to consider repurposing POC HIV VL tests as HIV VL self-tests. In the same way that people living with diabetes have reported that self-management has imparted increased empowerment and ownership of their condition, this could increasingly be true for people living with HIV.

HIV VL self-testing could be useful for a variety of purposes, especially given the increasing shift to remote care that we have seen in recent years, and in particular as a result of the COVID-19 pandemic. It could be of particular use for people living with HIV who are highly adherent, have been virally suppressed for a significant amount of time, are at low risk of developing drug-resistance, exhibit few treatment side effects, and do not require regular face-to-face meetings with their healthcare team. However, an annual clinic visit would likely still be necessary. This model may well be more affordable, require less clinician time and could be more acceptable tor people living with HIV than attending appointments in person.

## Methods

### Public consultation to understand needs of people living with HIV

As part of a wider patient and public involvement (PPI) programme, we held a consultation for people living with HIV in 2018 to better understand the needs and concerns of potential end users of such an intervention. PPI is a vital component of any health research, and we endeavoured to conduct the consultation early in our research programme, to allow the results to have maximum impact on our work developing HIV VL POC and self-testing devices. We followed guidance from INVOLVE, the UK national advisory group for the advancement and promotion of public involvement. According to INVOLVE, there is no requirement for ethical approval when undertaking PPI work^14^. We contacted the UCL Research Ethics Committee to confirm this.

### Recruitment and eligibility requirements of participants

Participants were recruited through Positive East, a London-based HIV charity. The participant eligibility requirements included (a) being over the age of 18, (b) ability to speak English, and (c) to have been on HIV ART for two years or more. Eighteen participants attended the consultation. All 18 satisfied the eligibility requirements and all signed a participant information and consent form upon arrival. At the end of the consultation, participants received an Amazon voucher and had their travel expenses reimbursed.

### Consultation structure

The format of the consultation involved short talks outlining a particular topic, followed by focus group discussions facilitated by researchers, finally concluded with a feedback session. The four sessions focused on (a) HIV VL measurements; (b) operational characteristics of a HIV VL self-test; (c) integrating HIV VL self-tests with digital healthcare technology; and (d) online data security of HIV VL self-tests. In addition, participants completed anonymous pre- and post-workshop surveys which included demographic questions, HIV clinic appointments and their thoughts on HIV VL testing.

### Data collection

We felt that video or audio recordings would not be acceptable for some participants so data was collected from the focus group discussions and feedback sessions in the form of flipchart mind mapping notes and post-it notes, which were written by workshop participants. Data was also available from the pre- and post-workshop surveys.

### Thematic analysis

All text from flipcharts and post-it notes was transcribed to form a data corpus for inductive thematic analysis. Individual flipcharts and post-it notes used in the focus groups were checked to ensure all content was included in the group codes. The data was somewhat already coded due to the brevity of the flipchart and post-it note statements. The data was coded individually by two coders who checked their individual datasets and discussed any discrepancies. It was not possible to code some statements, either because the meaning was ambiguous, or there was not a sufficient understanding of the meaning of the statement. Similarities and differences in coding approaches were discussed and overarching themes and sub-themes were agreed. Themes were then quantified according to how many times they were identified in each of the four sessions. These themes were then used to create word clouds, with the size of the words proportional to the number of times that theme was identified in a given session.

## Results

### Participant characteristics

The 18 participants were demographically diverse in terms of age (ranging from 25-74 years old, with 50% (n= 9/18) in the 45-54 years of age category), gender 61% (n= 11/18) women, ethnicity, sexual orientation, level of education and employment status (Table 1). All were over the age of 18 and had been on ART for two or more years.

**Table 1.** Characteristics of participants of the PPI consultation.

| Characteristic |  | N | % |
| --- | --- | --- | --- |
| **Age** | 25-34 years old | 1 | 5.6 |
|  | 35-44 years old | 3 | 16.7 |
|  | 45-54 years old | 9 | 50.0 |
|  | 55-64 years old | 4 | 22.2 |
|  | 65-74 years old | 1 | 5.6 |
| **Gender** | Male | 7 | 38.9 |
|  | Female | 11 | 61.1 |
| **Highest level of educational achievement** | Higher degree | 2 | 11.1 |
|  | Degree or equivalent | 7 | 38.9 |
|  | Higher education | 2 | 11.1 |
|  | A Level or equivalent | 2 | 11.1 |
|  | GCSEs grades A*-C | 2 | 11.1 |
|  | Other qualifications | 1 | 5.6 |
|  | No qualification | 1 | 5.6 |
|  | Prefer not to say | 1 | 5.6 |
| **Employment status** | Employed | 4 | 22.2 |
|  | Out of work and looking for work | 1 | 5.6 |
|  | Out of work, not currently looking for work | 3 | 16.7 |
|  | Retired | 2 | 11.1 |
|  | Unable to work | 6 | 33.3 |
|  | Prefer not to say | 2 | 11.1 |
| **Marital status** | Single, never married | 8 | 44.4 |
|  | Married/civil partnership | 1 | 5.6 |
|  | Widowed | 5 | 27.8 |
|  | Separated | 3 | 16.7 |
|  | Prefer not to say | 1 | 5.6 |
| **Ethnicity** | White British | 2 | 11.1 |
|  | White & Black Caribbean | 1 | 5.6 |
|  | White & Black African | 1 | 5.6 |
|  | White & Asian | 1 | 5.6 |
|  | Mixed British/Puerto Rican | 1 | 5.6 |
|  | Other Asian background | 1 | 5.6 |
|  | Black African | 9 | 50.0 |
|  | Black Caribbean | 2 | 11.1 |

### Results from surveys

We asked participants how often their HIV VL is currently tested. Three (17%) of 18 participants were tested every three months, 14 (78%) every six months, and one (6%) every 12 months. Most (72% or 13/18) participants were content with the frequency of their VL tests. There was one participant who wanted less frequent VL tests, and four wanted more frequent testing. Six (33%) participants reported that they kept their own record of their VL results, and a further eight (44%) said they currently did not, but would be interested in doing so. In the post-workshop survey, 14 (78%) of the 18 participants agreed or strongly agreed with the statement “*Monitoring my VL myself would feel empowering”* and 13 (72%) disagreed or strongly disagreed with the statement “*I don’t want the extra hassle of having to do it myself*”.

The session focusing on the operational characteristics of an HIV VL self-test included discussions around sample type and required volume, required time, accuracy, forms of readout and potential integration with a digital device for recording and communicating results to healthcare teams. One aspect not discussed was the cost of the test as it was beyond the remit of this consultation.

The accuracy of an HIV VL self-test is clearly of paramount importance. After a brief background discussion on sensitivity and specificity, we asked participants to discuss the importance of each. While the researchers had anticipated the importance of sensitivity (i.e. detecting the HIV virus even at very low levels) to participants, we had underestimated the significance to people living with HIV of specificity, and the effect a false positive result could have on the emotional wellbeing of the individual.

Participants voiced concerns over providing a fingerstick blood sample that is similar in volume to those typically required for sexually transmitted infections self-sampling kits (approximately 500 µL). Other anxieties included the storage and possible delivery of the tests, and their disposal.

We explored the possibility of a smartphone application storing a record of the individual’s VL tests, and this was of great interest to participants. While one third (6/18) participants already kept a record of their VL test results, there were eight (44%) who did not keep a record but would be interested in doing so, and further four (22%) who did not keep a record and were not interested in doing so.

### Results of the thematic analysis

The resulting themes from the thematic analysis of each of the four sessions was displayed in word clouds, with the more frequently identified themes appearing larger than the less frequently observed themes (Figure 1).

**Figure 1.**
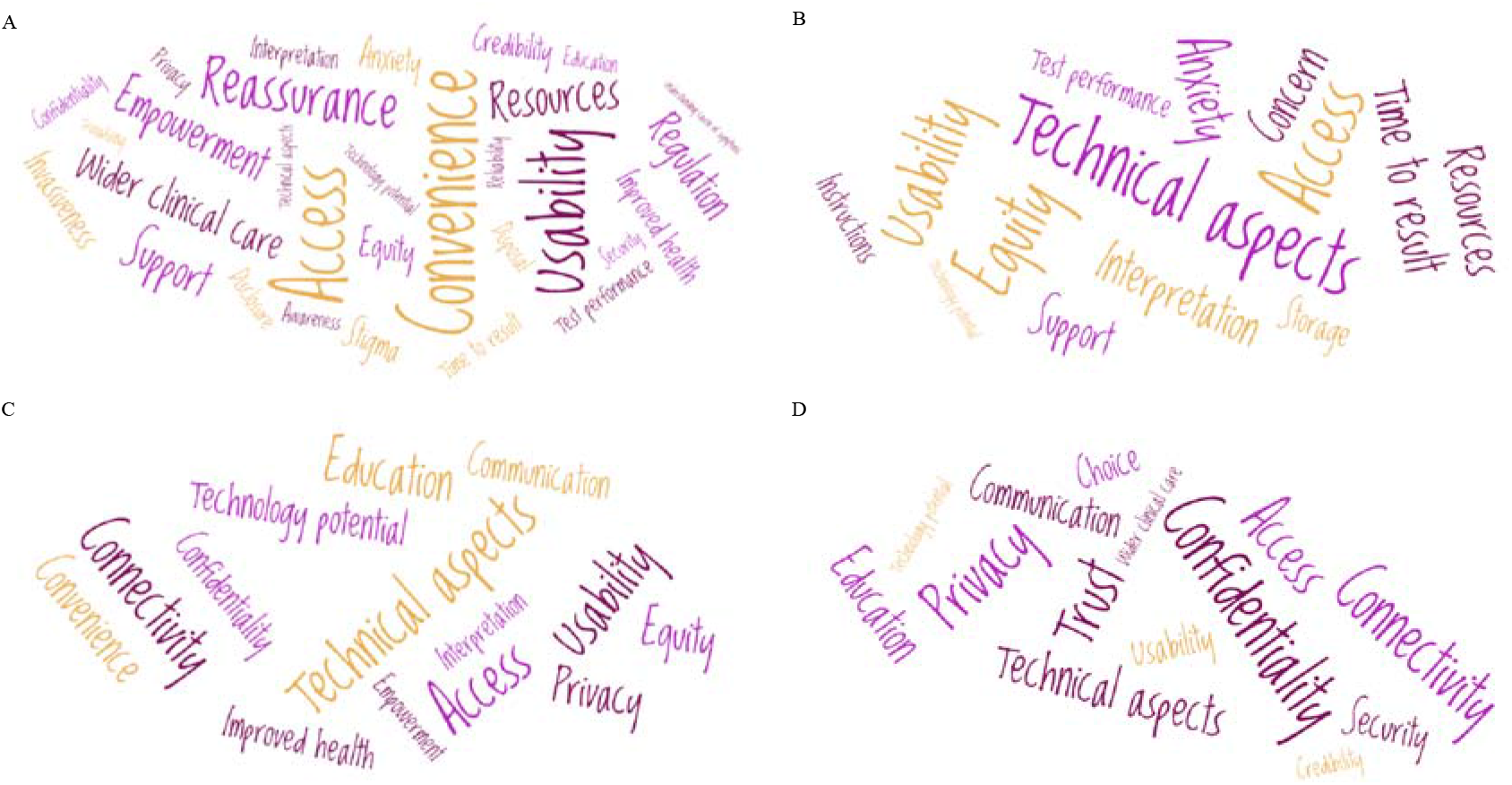
Resulting themes from the thematic analysis of data from each of the workshop sessions, which focused on (a) HIV VL measurements; (b) operational characteristics of an HIV VL self-test; (c) integrating HIV VL self-tests with digital healthcare technology; and (d) online data security of HIV VL self-tests. The size of the theme is proportional to the number of times it was referred to by participants during that session.

In our first session (Figure 1a), the key themes that emerged were related to individual responses to the prospect of HIV VL self-tests. Convenience, access and usability of the tests were the most prominent themes, but there were many references, both positive and negative, to potential impacts on their wider clinical care. Participants placed great value on their relationship with their clinical team, although many commented that some appointments were inconvenient, and reducing the number of visits to the clinical might be helpful. The second session (Figure 1b) focused on the operational characteristics of an HIV VL self-test, so it is not surprising that the two most conspicuous themes were technical aspects and usability. To simplify the analysis, we coded statements referring to accuracy, time to result, storage, disposal and invasiveness as technical aspects of the test. The technology of an HIV VL self-test is clearly fundamental to its uptake. It must be accurate but also easy to use, store, and dispose of. Some participants had concerns about the effects of having a self-test on accessibility and equity for some individuals. In the third session (Figure 1c), we introduced the potential for the test to be integrated with digital technology. The two major themes for this session were connectivity and technology potential. Some participants also had strong opinions that integration with digital technology should not hinder the access or equity of a potential test to members of society who are less familiar with digital technology. Our final session (Figure 1d) involved discussions around data security. The most prominent themes that emerged were confidentiality, privacy and trust. Stigma, communication and education were also discussed. Many of the workshop participants stated that since they already used internet banking on their smartphones with confidence, they would feel comfortable using HIV VL self-test in conjunction with their smartphone provided it had the same level of security.

## Discussion

This consultation is the first to explore the use of HIV VL self-testing in a UK context. While HIV VL self-testing is yet to become a reality, our PPI consultation allowed us to explore potential future challenges and will help shape our research in this space. It enabled us to consider attitudes towards the current practice of VL testing and digital healthcare technologies, and explore how these may affect the acceptability of HIV VL self-testing in the future. The small group discussions gave insights into the importance of various technical aspects of the test, including requirements for high specificity and ease-of-use.

One key limitation of our thematic analysis is that the data corpus we used was typically in the form of short statements, which is not a traditional form of data for thematic analysis. It is possible that some of the richness and depth of data was lost due to the brevity of the statements. However, recording data from small group discussions in this form was considered more acceptable to attendees than audio recording. Future work could include in-depth interviewers with potential end users in order to further understand attitudes towards HIV VL self-testing. Another limitation is the small sample of individuals we consulted, since this was an early stage PPI workshop. Future work will need a larger sample size to provide more reliability of the results, and ensure they are truly representative of the views of people living with HIV.

Implementing new health technologies is complex. It requires thorough consultation with end users in order to ensure successful adoption, scale up and long-term sustainability. However, the increase in remote consultations that has been brought about by COVID-19 has demonstrated that new health technologies can be successfully adopted when there is a clear rationale and well-designed solution. It is possible that the HIV care continuum may change and adapt in future years, and particularly as we look beyond the COVID-19 pandemic. Current POC VL tests require further development in order to reach a stage where they can be used by the individual as self-tests. Once this is achieved, robust analyses of accuracy, feasibility, acceptability and cost-effectiveness will be required, as well as the necessary funding and regulatory approval, in order to make HIV VL self-testing a reality.

## Data Availability

All data is available in the Supplemental Material.

## Author Contributions

Conceptualization: Harriet D Gliddon, Rachel A McKendry, Jo Gibbs; Methodology: Harriet D Gliddon, Jo Gibbs; Formal analysis and investigation: Harriet D Gliddon, Rachel A McKendry, Jo Gibbs; Writing - original draft preparation: Harriet D Gliddon, Jo Gibbs; Writing - review and editing: Harriet D Gliddon, Rachel A McKendry, Jo Gibbs; Funding acquisition: Harriet D Gliddon, Rachel A McKendry, Jo Gibbs; Resources: Harriet D Gliddon, Rachel A McKendry, Jo Gibbs; Supervision: Rachel A McKendry, Jo Gibbs.

## Acknowledgements

The authors wish to thank all who attended the workshop and facilitated sessions, and Positive East who helped with the facilitation and logistics. HDG and RAM are funded by the i-sense EPSRC IRC in Early Warning Sensing Systems in Infectious Disease (EP/K031953/1) and i-sense: EPSRC IRC Next Steps Award in Agile Early Warning Sensing Systems for Infectious Diseases and Antimicrobial Resistance (EP/R00529X/1). RAM acknowledges the Royal Society Wolfson Research Merit Award. The workshop was funded by a UCLH NIHR BRC Patient and Public Involvement Starter Grant.

